# Transmission of SARS-CoV-2 before and after symptom onset: impact of nonpharmaceutical interventions in China

**DOI:** 10.1101/2020.12.16.20214106

**Authors:** Mary Bushman, Colin Worby, Hsiao-Han Chang, Moritz Kraemer, William P. Hanage

## Abstract

Nonpharmaceutical interventions, such as contact tracing and quarantine, are currently the primary means of controlling the spread of SARS-CoV-2; however, it remains uncertain which interventions are most effective at reducing transmission at the population level. Using serial interval data from before and after the rollout of nonpharmaceutical interventions in China, we estimate that the relative frequency of presymptomatic transmission increased from 34% before the rollout to 71% afterward. The shift touward earlier transmission indicates a disproportionate reduction in transmission post-symptom onset. We estimate that, following the rollout of nonpharmaceutical interventions, transmission post-symptom onset was reduced by 82% whereas presymptomatic transmission decreased by only 16%. These findings suggest that interventions which limit opportunities for transmission in the later stages of infection, such as contact tracing and isolation, may have been particularly effective at reducing transmission of SARS-CoV-2.

## Introduction

In January 2020, in Wuhan, China, what began as a cluster of viral pneumonia cases rapidly spiraled into an epidemic of a new disease, COVID-19, caused by a novel coronavirus, designated SARS-CoV-2. As cases mounted in Wuhan and began cropping up elsewhere, China introduced a comprehensive set of measures, termed nonpharmaceutical interventions (NPIs), to contain the virus. On January 23, a lockdown was enacted in Wuhan, which shut down public transit and travel out of the city. Other cities throughout Hubei province (of which Wuhan is capital) announced similar lockdowns over the next few days [1]. The rest of China was subject to social distancing measures: mass transit and public gatherings were severely curtailed, and the New Year holiday (*Chunyun)* was extended, which kept most schools, workplaces, and businesses closed [1-3]. In addition, numerous measures were implemented to rapidly identify and isolate suspected cases. These included temperature checks at borders and travel hubs, quarantine of new arrivals, isolation of both confirmed and suspected cases, and contact tracing with quarantine and medical observation.

China’s robust public health response was decidedly effective in controlling the spread of SARS-CoV-2 [4-7]. As of December 1, 2020, China had a cumulative incidence of 63 cases per million residents and cumulative mortality of 3 deaths per million, compared to 7937 cases and 186 deaths per million persons globally [8]. Given the magnitude and intensity of the response in China, which would be difficult to replicate in many settings, it would be useful to know which interventions were most effective in limiting the spread of the virus.

Evaluating the effectiveness of specific NPIs has been a challenge throughout the COVID-19 pandemic because multiple interventions are typically used concurrently. Several studies have estimated the combined impact of multiple NPIs [5-7, 9]; others have estimated the effect of specific control measures using statistical approaches [10-20] or modeling [21-25]. The majority of studies focus on the effectiveness of social distancing interventions, while relatively few explicitly consider the impact of quarantine or isolation measures. However, in China, the impact of isolation has been indirectly observed in a shift toward earlier transmission, reflected in shorter serial intervals [26]. Here, we use serial interval data and incidence data from China to show that, in the first few weeks after NPIs were implemented, presymptomatic transmission decreased slightly but transmission post-symptom onset declined dramatically. These results suggest that “post-onset” interventions, such as case isolation, may have been more effective than other control measures at limiting transmission of SARS-CoV-2.

## Results

We compiled published data, including symptom onset dates, for 873 infector-infectee pairs from China (see Methods) [27-30]. Among these case pairs, the majority of transmission events occurred outside Hubei, with 84% of secondary cases acquired in other provinces (Table S1). Symptom onset dates ranged from January 7 to February 29, 2020, a period that spans the rollout of nonpharmaceutical interventions in China. The mean serial interval, defined as the time between onset of symptoms in infector and infectee, was 4.64 days over the entire period, similar to published estimates [27, 28, 31, 32], but this distribution shifted markedly over time. A linear regression of serial intervals vs. symptom onset dates of primary cases reveals a significant decrease in the length of serial intervals (p = 4×10^−34^, Figure 1a), an observation also made by Ali et al. [26].

**Fig. 1.**
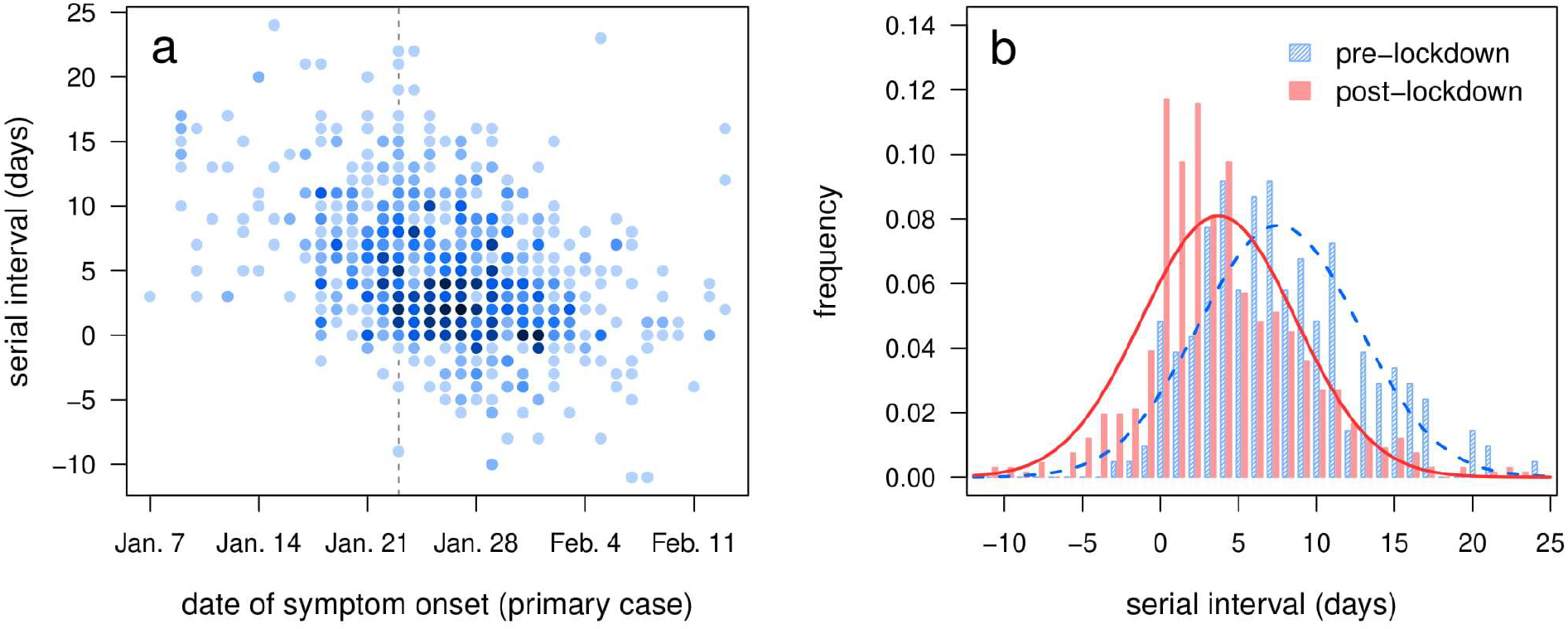
Serial intervals of case pairs in China. (a) Serial interval vs. date of symptom onset in the primary case of each pair. Shading reflects the number of overlapping points, with darker colors indicating higher numbers. Dashed vertical line indicatesJan 23, the date of the initial lockdown in Wuhan. (b) Serial interval histograms and best-fit normal distributions for primary cases with symptom onset before Jan 23 (pre-lockdown; blue, hatched bars and dashed line) and on/after Jan 23 (post-lockdown; red, solid bars and solid line).

We then divided case pairs into two time periods using the date of symptom onset in each primary case. Those with onset before January 23 - marking the lockdown of Wuhan and the start of a national rollout of nonpharmaceutical interventions – were denoted pre-lockdown (n = 207) and the rest denoted post-lockdown (n = 666). Pre- and post-lockdown serial intervals were significantly different between the two time periods (p = 4×10^−19^, Figure 1b), with a mean of 7.57 days (standard deviation 5.13 days) before the lockdown and a mean of 3.73 days (standard deviation 4.93 days) afterward.

Next, we used a Markov chain Monte Carlo (MCMC) approach to estimate the distribution of the generation interval for each time period by fitting to serial interval data. We replicated this analysis using three different priors for the incubation period distribution (Table S2), based on work by Lauer et al. [33], Zhang et al. [30]; and Backer et al. [34]; we also used two different models for the generation interval distribution. Under the incubation-independent model, all generation intervals are drawn from a single gamma distribution; under the incubation-dependent model, the generation interval is still gamma-distributed but the probability distribution for each case is horizontally “stretched” in proportion to the incubation period, which means that cases with longer incubation periods will tend to have longer generation intervals.

We used a modified deviance information criterion [35] to assess model fit (S1 Text) and found that the best model for both time periods was the incubation-independent model – which, somewhat surprisingly, suggests generation intervals do not increase with incubation period - paired with the prior based on Lauer et al. (Table S3).

As expected, the fitted generation interval distributions had similar means but smaller variances than the serial interval distributions [36]. Under the best model, pre-lockdown generation intervals had a mean of 7.50 days (standard deviation 3.95 days) while post-lockdown generation intervals had a mean of 3.90 days (standard deviation 3.15 days). Under the remaining models, mean generation intervals ranged from 7.49 to 7.64 days for the pre-lockdown period and from 3.84 to 4.04 days for the post-lockdown period (Table S4).

Using the inferred generation interval distributions, we estimated the relative frequency of presymptomatic transmission under each model by calculating the probability of the generation interval being shorter than the incubation period. Under the best model, the frequency of presymptomatic transmission was estimated to be 34.4% in the pre-lockdown period (95% credible interval 28.3% - 41.3%) and 71.0% in the post-lockdown period (95% CI 67.6% - 74.2%) under the best model (Figure 2). Across all models, the estimated frequency of presymptomatic transmission ranged from 30.7% to 47.0% for the pre-lockdown period, with 95% CIs collectively extending from 24.0% to 53.7% (Table S5). For the post-lockdown period, estimates ranged from 68.1% to 80.6%, with 95% CIs extending from 64.5% to 83.5%. We note that the estimates for the pre-lockdown period are lower than many published estimates of the frequency of presymptomatic transmission [27, 29, 37-40], which raises the possibility that many estimates might be inflated by the effects of nonpharmaceutical interventions.

**Fig. 2.**
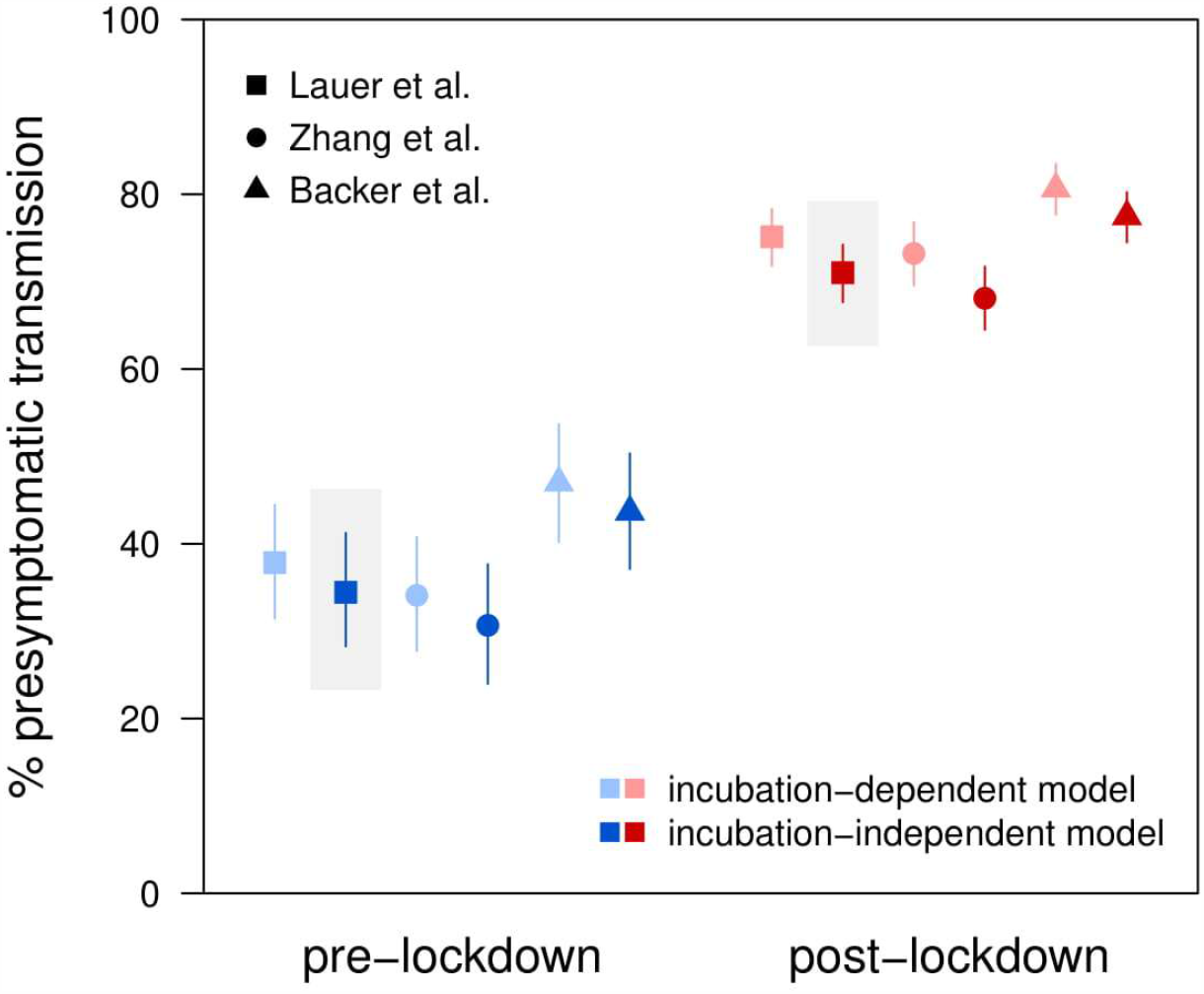
Estimated relative frequency of presymptomatic transmission before and after the first lockdown on January 23. Points and lines show posterior means and 95% credible intervals, respectively. Colors denote time periods (blue, pre-lockdown; red, post-lockdown), shades denote generation interval distribution models (light, incubation-dependent model; dark, incubation-independent model), and symbols denote sources for incubation period prior distributions (squares, Lauer et al. [33]; circles, Zhang et al. [30]; triangles, Backer et al. [34]). Estimates from the best model are highlighted in gray.

The shift toward presymptomatic transmission following the rollout of NPIs suggests a disproportionate reduction in transmission post-symptom onset. We therefore estimated the change in transmission during each phase of the infection (before and after symptom onset) following the implementation of control measures. To do so, it was first necessary to estimate the total reduction in transmission of SARS-CoV-2. For this, we used province-level incidence data for all of China, combining the numbers for all provinces except Hubei because the transmission events represented in the case-pair data mostly occurred outside Hubei. We calculated the daily case reproduction number (*R*_*t*_), shown in Figure 3, using a version of the Wallinga-Teunis method [41] which was modified to allow for time-varying serial intervals, as changes in the serial interval distribution have been shown to affect inference of reproduction numbers [26].

**Fig. 3.**
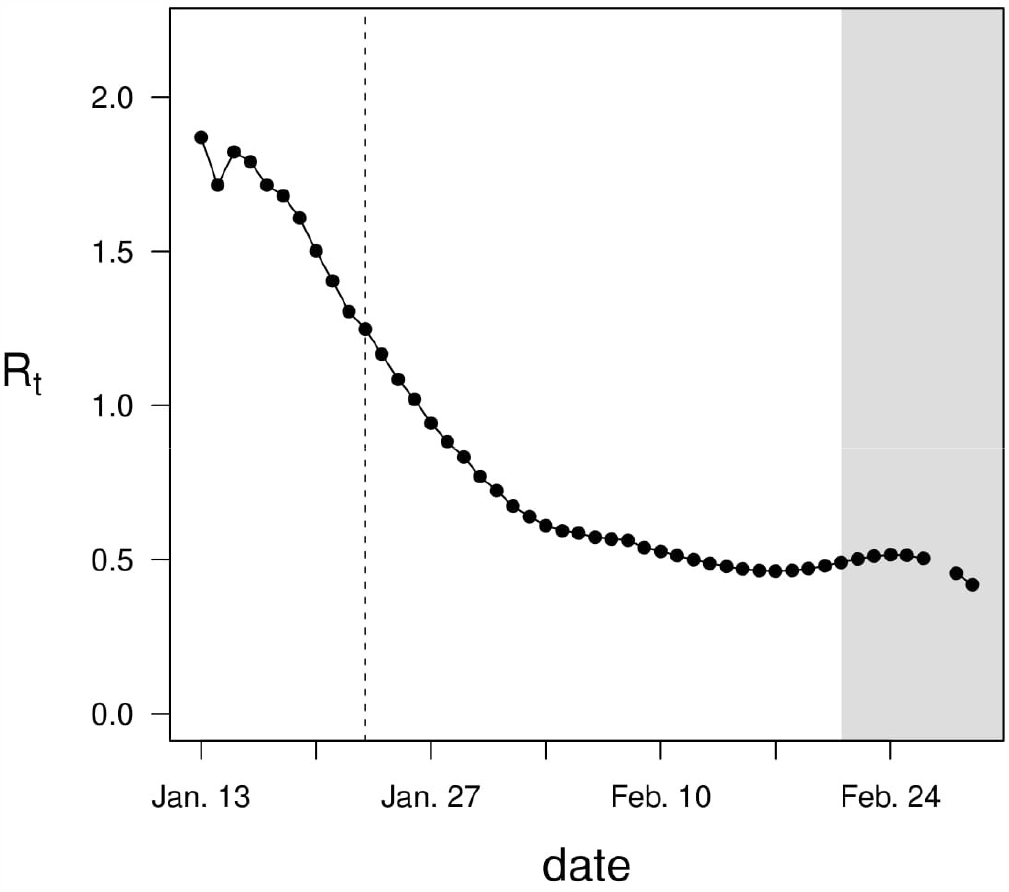
Daily reproduction numbers estimated using incidence data from all Chinese provinces except Hubei. Dashed vertical line, start of initial lockdown in Wuhan on Jan. 23; gray shading, region in which *R*_*t*_ is likely to be underestimated due to right-truncation.

We used the estimated *R*_*t*_ values to calculate the mean reproduction numbers for the pre- and post-lockdown periods; estimates past February 20 were not used due to the potential for underestimation of *R*_*t*_ as a result of right-truncation (Figure 3) [42]. The mean reproduction numbers were estimated to be 1.64 for the pre-lockdown period and 0.666 for the post-lockdown period, corresponding to a 59.4% reduction in overall transmission. Treating this net change as a weighted average of the changes in presymptomatic transmission and transmission post-symptom onset, we calculated the change in the absolute frequency of transmission during each phase of the infection. Under the best model, we estimate that presymptomatic transmission decreased by 15.5% (95% CI: −30.6% to +2.56%) while transmission post-symptom onset decreased by 82.0% (95% CI: −84.5% to −79.0%; Figure 4). Across all models, estimates of the change in presymptomatic transmission ranged from −29.9% to −8.62%, with 95% CIs extending from −39.4% to +16.3% (Table S6). Estimates of the change in transmission post-symptom onset ranged from −85.1% to −81.3%, with 95% CIs extending from −87.9% to −78.3%.

**Fig. 4.**
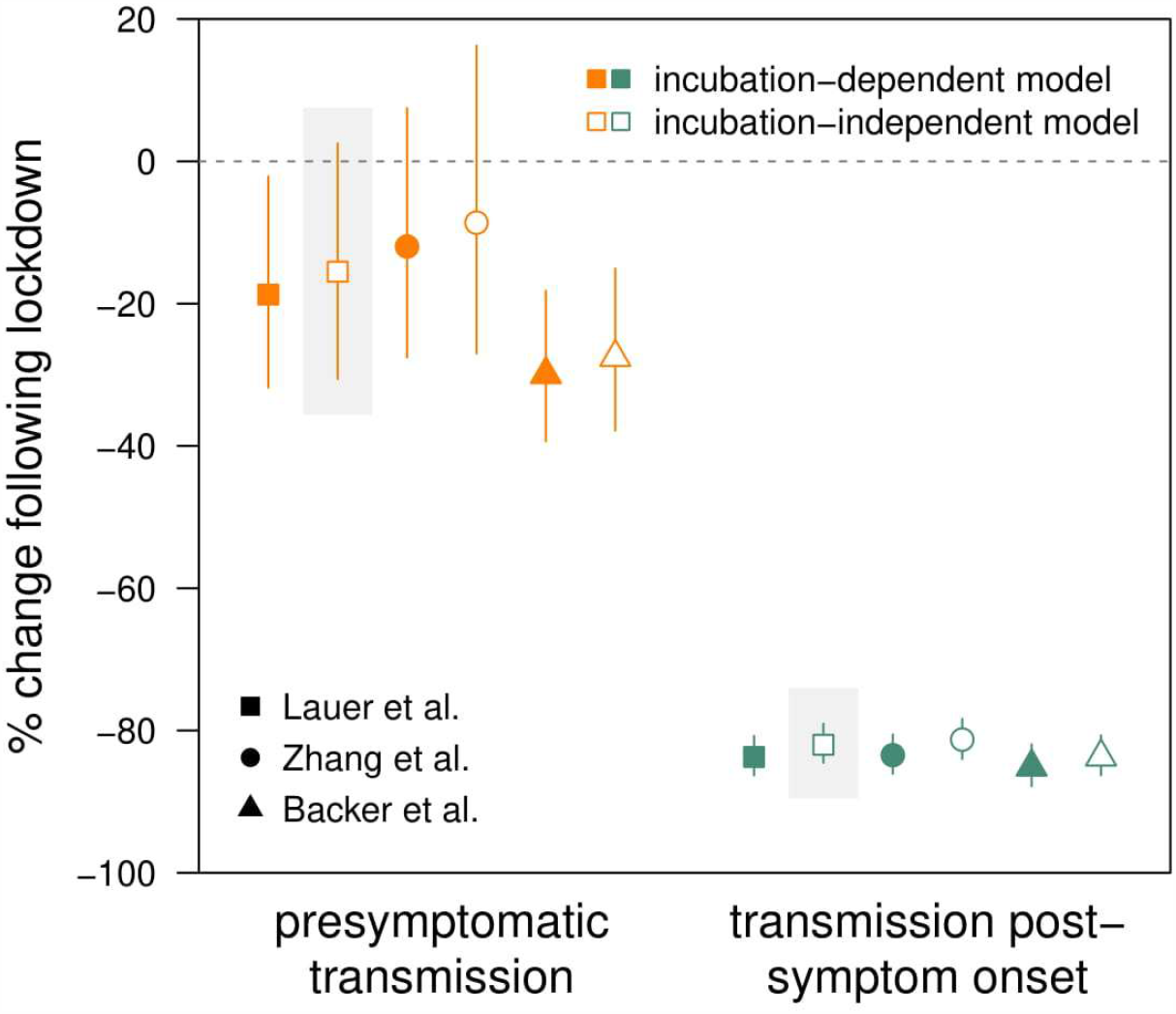
Estimated percent change in absolute frequency of presymptomatic transmission and transmission post-symptom onset. Points and lines show posterior means and 95% credible intervals, respectively. Orange, presymptomatic transmission; green, transmission post-symptom onset; closed symbols, incubation-dependent model; open symbols, incubation-independent model. Symbol shapes denote sources for incubation period prior distributions (squares, Lauer et al. [33]; circles, Zhang et al. [30]; triangles, Backer et al. [34]). Estimates from the best model are highlighted in gray.

One hypothesis to explain the disproportionate reduction in transmission post-symptom onset is that “post-onset” interventions, such as case isolation, were highly effective at preventing transmission in the later stages of infection; however, several alternative explanations must be considered. One possibility is that travel restrictions reduced the frequency of case pairs with different infection locations (primary and secondary cases infected in different cities), which might be expected to have longer-than-average serial intervals. Although the proportion of case pairs infected in different cities decreased after the lockdown (p=1×10^−4^; Table S7), a two-way ANOVA found no significant effect of location matching on serial intervals (p>0.05).

Another possibility is that social distancing increased time spent at home, with more frequent exposure leading to earlier transmission between family members or household contacts – similar to the relationship between force of infection and age of first infection. The proportion of transmission events occurring between family members increased following the lockdown (p=3×10^−5^), although there was no change in the proportion of transmission events taking place within households (p=0.09; Table S7). However, neither familial relationship nor household contact had a significant effect on serial intervals, nor did either factor have a significant interaction with time period (p>0.05).

## Discussion

Following the implementation of nonpharmaceutical interventions in China, the transmission of SARS-CoV-2 changed in two ways. Compared to the period preceding the lockdown of Wuhan on January 23, 2020, the post-lockdown period was characterized by a significant reduction in the reproduction number (*R*_*t*_), indicating decreased transmission, and a decrease in the length of generation intervals, reflecting earlier transmission. Specifically, we found that overall transmission of SARS-CoV-2 declined by 59.4% after the lockdown, while presymptomatic transmission expanded from 34% to 71% of all transmission events. The shift toward earlier transmission implies a disproportionate reduction in transmission following symptom onset; we estimate that presymptomatic transmission decreased by roughly 16% after the lockdown, whereas transmission post-symptom onset decreased by approximately 82%.

Several factors might contribute to the observed shift toward earlier transmission: a reduction in the proportion of primary cases imported from other cities, eliminating a potential delay in transmission; increased time at home, resulting in more frequent exposure to infectious household contacts; and/or interventions that disproportionately reduce transmission in the later stages of infection. Our results do not support either of the first two possibilities: whether a linked pair of cases were infected in the same city was not a significant predictor of the serial interval, nor was the existence of a shared household or familial relationship. The third possibility is indirectly supported by observed correlations between isolation delays and serial intervals, which suggest that case isolation is capable of shifting the serial interval distribution [25, 26, 43].

Our findings therefore suggest that “post-onset” interventions, such as case isolation, may have been responsible for the dramatic reduction in transmisson post-symptom onset that followed the implementation of NPIs in China. If so, this would indicate that isolation and associated interventions, such as contact tracing, were among the largest contributors to the rapid reduction in transmission of SARS-CoV-2, while the comparatively modest reduction in presymptomatic transmission would indicate a relatively small impact of social distancing measures. However, it is possible that the effects of social distancing were offset by increased household transmission, an effect that might be expected to dissipate once most household transmission chains go extinct.

A key limitation of this work is the lack of information regarding transmission from asymptomatic infections, which may comprise 40-45% of all SARS-CoV-2 infections [44]. Because the serial interval is defined by symptom onset dates, alternative methods are required to examine the impact of NPIs on asymptomatic transmission. For instance, data on exposure windows could be used to estimate the time of infection for primary cases, similar to the approach used for estimating incubation periods [30, 33, 34, 45, 46]; in conjunction with exposure or symptom onset dates for secondary cases, an approach similar to the one employed here could then be used to infer generation intervals for transmission pairs with asymptomatic primary cases.

In summary, we find that the implementation of nonpharmaceutical interventions in China was followed not only by a rapid decrease in the rate of SARS-CoV-2 transmission, but a significant shift in the timing of viral transmission, with more transmission occurring in the presymptomatic (incubation) period. The leading hypothesis to explain these observations is that interventions, particularly case isolation, were highly effective in limiting transmission in the later stages of infection, while other measures, such as social distancing, had a more limited impact on transmission in the earlier stages. These findings suggest that rapid case detection and isolation, if rigorously implemented, may be a highly effective strategy for interrupting transmission of SARS-CoV-2.

## Methods

### Data sources

We obtained data on serial intervals for linked pairs of cases reported in China in January and February 2020. We combined data that were previously collected and published by the following sources: Xu et al. with 679 case pairs, compiled from provincial and urban health commission reports [27]; Du et al. with 468 case pairs, compiled from provincial health agency reports [28]; He et al. with 41 case pairs, compiled from government and media reports [29]; and Zhang et al. with 35 case pairs, compiled from health agency and media reports [30]. We cross-checked all datasets and eliminated duplicate case pairs, which we identified as those with matching sex, age, and symptom onset date for both cases. This left a total of 873 unique case pairs for use in parameter estimation. Among these, a majority of transmission events took place outside Hubei province (home to the city of Wuhan), with 84.2% of secondary cases infected outside Hubei (Table S1).

### Analysis of serial intervals

Since dates of transmission were unknown, we dated each serial interval using the date of symptom onset in the primary case. We used simple linear regression (serial interval versus date) to test the hypothesis that serial intervals declined over time. We then divided serial intervals into two time periods based on the date of symptom onset in the primary case of each pair. Case pairs were designated “pre-lockdown” if the primary case developed symptoms before the lockdown of Wuhan on Jan 23 (i.e. symptom onset occurred on or before Jan 22) and as “post-lockdown” otherwise. The terms pre-lockdown and post-lockdown refer here to the time periods preceding and following a specific event – the lockdown of Wuhan – rather than periods characterized by the presence or absence of lockdown measures; true lockdowns were not widely implemented beyond Hubei province. We used a two-sided Student’s t-test to test the hypothesis that the mean serial intervals differed between the pre- and post-lockdown periods.

### Generation interval distributions

Consider a linked pair of cases, with the secondary case arising by transmission from the primary case. We denote the incubation periods of the primary and secondary cases by *θ*_1_ and *θ*_2_, respectively. We use *δ* to refer the serial interval and *τ* to denote the generation interval. The relationship between these quantities is given by *τ* = *δ* + *θ*_1_ – *θ*_2_ (Figure 5).

**Fig. 5.**
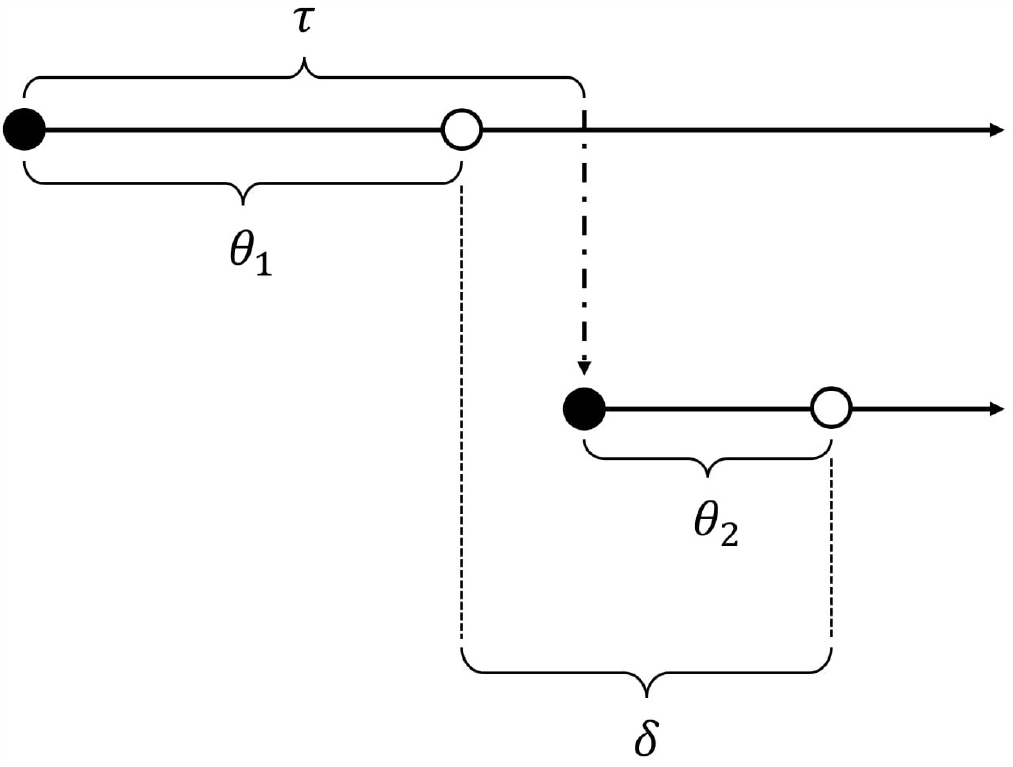
Diagram showing incubation period (*θ*), serial interval (*δ*), and generation interval (*τ*) for a linked pair of cases. Closed circle, time of infection; open circle, time of symptom onset; dot-dash arrow, transmission from primary case to secondary case.

We assumed that the generation interval *τ* followed a gamma distribution *f*_*τ*_ with unknown parameters *α* and *β*. Due to uncertainty regarding the relationship between the incubation period and the generation interval, we used two different models for the generation interval distribution. In the incubation-independent model, we assumed that a single generation interval distribution, with shape parameter *α* and rate parameter *β*, applied to all individuals, regardless of incubation period. In the “incubation-dependent” model, we assumed that individuals with longer incubation periods would tend to have longer generation intervals; specifically, for a primary case with incubation period *θ*_1_, we assumed that the generation interval followed a gamma distribution with shape parameter *α* and rate parameter *β*/*θ*_1_. This is equivalent to defining a new random variable *X* = *τ*/*θ*_1_ where *X* follows a gamma distribution with shape *α* and rate *β*. This formulation causes the generation interval distribution to be horizontally “stretched out” in proportion to *θ*_1_ – for instance, it results in the expected generation interval being a fixed multiple of *θ*_1_, rather than a fixed length of time.

### Parameter estimation

We used a Markov chain Monte Carlo (MCMC) algorithm to estimate the parameters of the pre- and post-lockdown generation interval distributions by fitting to serial interval data from each time period. Because an observed serial interval depends on the incubation period of each infection as well as the generation interval, we also estimated the unobserved incubation periods *θ*_1_ and *θ*_2_ for each pair of cases (data augmentation). The model assumed that the incubation periods were drawn from a prior *f*_*θ*_; we used three different priors drawn from the literature (see below). The generation interval was assumed to be drawn from a gamma distribution *f*_*τ*_ with unknown parameters *α* and *β*, with minimally informative priors *f*_*α*_ and *f*_*β*_, respectively (Table S2). We can express the joint posterior density of the unknown parameters as follows: 

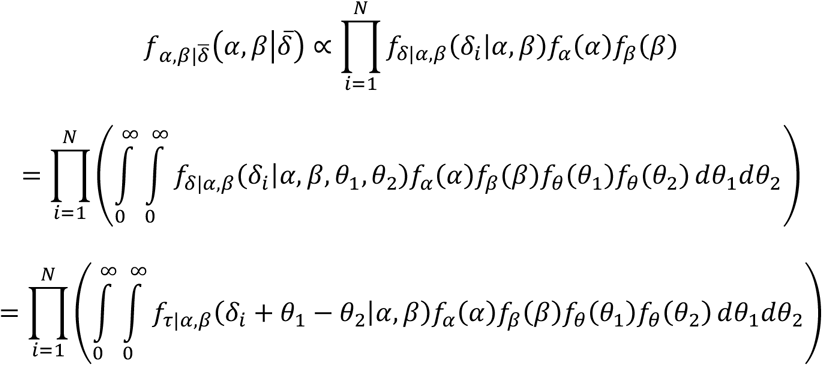

We estimated the unknown quantities using a Metropolis-Hastings algorithm, with each iteration taking place in two parts. The parameters *α* and *β* were updated first, with proposed values *α*′ and *β*′ being accepted with probability min 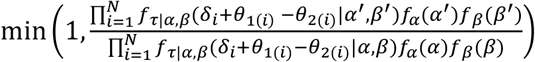. The incubation periods *θ*_1_ and *θ*_2_ for each case pair were then updated, with proposed values *θ*_1(*i*)_′ and *θ*_2(*i*)_′ being accepted with probability min 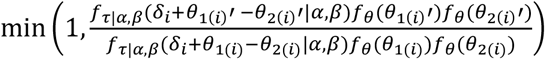.

We ran the algorithm for 250,000 iterations, discarded the first 50,000 and thinned the remainder by keeping every 10^th^ iteration. The resulting set of 20,000 observations was used to approximate the joint posterior distribution of the parameters of interest. The aggregated posterior distributions of the incubation periods *θ*_1_ and *θ*_2_ for each model are illustrated in Figures S1-S4, while posterior means and 95% credible intervals for the generation interval parameters are reported in Tables S3-S4. We report the mean and standard deviation of the generation interval distribution alongside the parameters *α* and *β*, since these latter quantities are difficult to interpret. For the incubation-independent model, the mean and variance are simply *α*/*β* and *α*/*β*^2^, respectively. For the incubation-dependent model, the mean generation interval is given by E[*X*]E[*θ*], where *X* = *τ*/*θ* follows a gamma distribution with shape *α* and rate *β*. The variance is given by (Var[*X*] + E[*X*]^2^)(Var[*θ*] + E[*θ*]^2^) – (E[*X*]^2^)(E[*θ*]^2^). We calculated the mean and variance of the generation interval for each iteration in the converged and thinned Markov chain in order to approximate posterior distributions, which we used to obtain posterior means and 95% credible intervals.

### Incubation period distributions

Since the incubation period distribution affects the inferred generation interval distribution, we replicated our analysis using three different priors (*f*_*θ*_) for the incubation period. The distributions and their sources are as follows: a lognormal distribution with mean 5.52 days and standard deviation 2.41 days, based on Lauer et al. [33]; a lognormal distribution with mean 5.21 days and standard deviation 2.59 days, based on Zhang et al. [30]; and a Weibull distribution with mean 6.49 days and standard deviation 2.35 days, based on Backer et al. [34]. The parameters for these distributions can be found in Table S2.

### Model fit

We assessed model fit using a modified deviance information criterion (DIC) for data-augmented models (S1 Text) [35]. Broadly speaking, DIC is a generalization of the Akaike information criterion (AIC); it penalizes model complexity as well as poor model fit (low likelihood of observing the data under the specified model). As with AIC, the “best” model is the one with the lowest DIC value.

### Pre-symptomatic transmission

We next used the joint posterior distribution of the generation interval parameters to approximate the distribution of the percentage of transmission expected to take place prior to symptom onset, which we denote *φ*. For the incubation-independent model, given *α* and *β*, the proportion of transmission expected to occur before symptom onset is given by 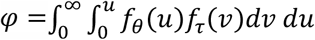, where *f*_*τ*_ is a gamma distribution with shape *α* and rate *β*. For the incubation-dependent model, the expected proportion of transmission occurring before symptom onset is given by 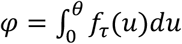, where *f*_*τ*_ is a gamma distribution with shape *α* and rate *β*/*θ*. We calculated *φ* for each iteration in the converged and thinned Markov chain in order to approximate the posterior distribution of *φ*, which we use to obtain posterior means and 95% credible intervals.

### Method to estimate reproduction numbers (R_t_) with time-varying serial intervals

We introduce a modified version of the Wallinga-Teunis method of estimating the reproduction number at time *t* (denoted *R*_*t*_). In the basic method described by Wallinga & Teunis (2004), the probability that case *i* was infected by case *j* is given by 

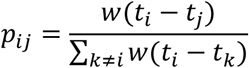

where *t*_*i*_ is the time at which case *i* developed symptoms, and *w* the serial interval distribution. The reproduction number for case *j* is therefore given by 

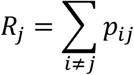

and the reproduction number at time *t* (*R*_*t*_) is equal to the reproduction number for any case with symptom onset at time *t*.

We now present a modification of this approach, which allows for time-varying serial interval distributions. Rather than a fixed serial interval distribution *w*, let *w*_*t*_ be the distribution of serial intervals for primary cases with symptom onset in the time window [*t* – *z, t* + *z*]. Then the probability case *i* was infected by case *j* is given by 

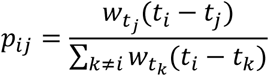

and the reproduction number for case *j* is similarly given by *R*_*j*_ = ∑_*i ≠j*_ *p*_*ij*_, which we can rewrite as follows: 

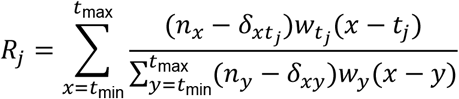

where *t*_min_ and *t*_max_ are the first and last dates of symptom onset, *n*_*t*_ is the number of cases with symptom onset at time *t*, and *δ*_*ij*_ is the Kronecker delta function, with 

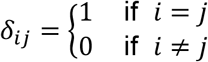

such that case *j* is subtracted from the set of potential infectees with symptom onset at time *t*_*j*_. With *R*_*t*_ assumed to be equal to the reproduction number for any case with symptom onset at time *t*, we can modify the above to get the final expression for *R*_*t*_: 

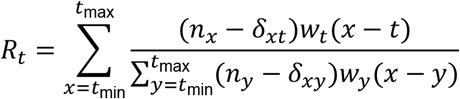

### Estimation of time-varying serial interval distributions

We used the serial interval data described above to estimate the time-varying serial interval distributions; we assumed serial intervals for primary cases with symptom onset at time *t* followed a normal distribution with mean and standard deviation calculated using serial intervals for primary cases with symptom onset in the window [*t* – *z, t* + *z*] with *z* = 3 (corresponding to a 7-day moving window). For time windows extending beyond the range of primary case symptom onset dates, the nearest 7-day window falling within this range was used to estimate the serial interval distribution.

### Estimation of reproduction number (R_t_) using case incidence data

The incidence data based on reported case pairs are less than ideal for estimation of *R*_*t*_ due to small daily case numbers (median of 9 cases per day) as well as potential sampling biases. We therefore compared case pair incidence data to the incidence data for all of China minus Hubei province. Incidence curves for the two datasets were similar, although the non-Hubei curve was shifted to the right (Figure S5), presumably due to the delay between symptom onset and case reporting (nationwide data did not include symptom onset dates). After shifting the non-Hubei incidence data back by 7 days to align the peaks of the two curves, we find good agreement between the case pair data and non-Hubei data (Figure S6).

We estimated *R*_*t*_ using both the case pair incidence data and the time-shifted non-Hubei incidence data, and find good agreement between the two sets of estimates (Figure S7). However, because the non-Hubei data feature larger case numbers, and presumably smaller sampling error, we use the *R*_*t*_ estimates based on the these data for subsequent analysis.

### Estimating mean reproduction numbers before and after lockdown

We used *R*_*t*_ estimates from before and after the lockdown (before Jan. 23 and on/after Jan. 23, respectively) to estimate the mean reproduction numbers for the pre- and post-lockdown periods. In order to reduce the effects of sampling error, we only used *R*_*t*_ estimates for days with at least 10 cases. In addition, because the Wallinga-Teunis method uses future case incidence to estimate the reproduction number, it will tend to underestimate *R*_*t*_ toward the end of a time series due to right-truncation. We therefore calculated the probability of a secondary case developing symptoms by *t*_max_ if the primary case developed symptoms at time *t*, given the serial interval distribution at time *t* (Figure S8). We discarded *R*_*t*_ estimates beyond the time point at which this probability dropped below 90%.

### Estimating reductions in presymptomatic and (post)symptomatic transmission

Let *R*_pre_ and *R*_post_ denote the mean reproduction numbers for the prelockdown and postlockdown periods, respectively. Similarly, let *φ*_pre_ and *φ*_post_ denote the relative frequency of presymptomatic transmission for each time period. Then the change in the absolute frequency of presymptomatic transmission (from prelockdown to postlockdown) is given by 

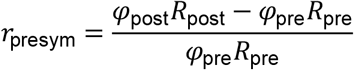

and the change in the absolute frequency of (post)symptomatic transmission is given by 

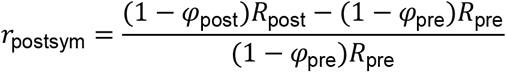

Posterior distributions for the changes in presymptomatic and (post)symptomatic transmission were obtained using the posterior distributions for the relative frequency of presymptomatic transmission for each time period.

### Software

All of the analysis for this study was conducted in R (version 3.6.1).

## Data Availability

All data are freely available through Open Science Framework.

https://osf.io/ezmkq/

## Declarations

### Funding

This work was supported by the National Institute of Allergy and Infectious Diseases (R01AI128344).

### Competing interests

The authors declare that no competing interests exist.

### Ethical approval

Because the data utilized for the current project were not collected specifically for the present study and because no one on the study team had access to identifying data, the study was not considered human subjects research and a Harvard Longwood Campus Institutional Review Board application was not required.

### Consent to participate

Not applicable

### Consent for publication

Not applicable

### Availability of data and material

The serial interval and incidence data are freely available through Open Science Framework (DOI: 10.17605/OSF.IO/EZMKQ).

### Code availability

The code files and model output are freely available through Open Science Framework (DOI: 10.17605/OSF.IO/EZMKQ).

### Author contributions

All authors contributed to study conception and design. MB, CJW and HHC developed the methodology. MB analyzed the data and drafted the manuscript. All authors revised the manuscript and approved the final version.

## Notes

### Competing Interest Statement

The authors have declared no competing interest.

