## Supplementary material for "Transmission of SARS-CoV-2 before and after symptom onset: impact of nonpharmaceutical interventions in China": Fig S4

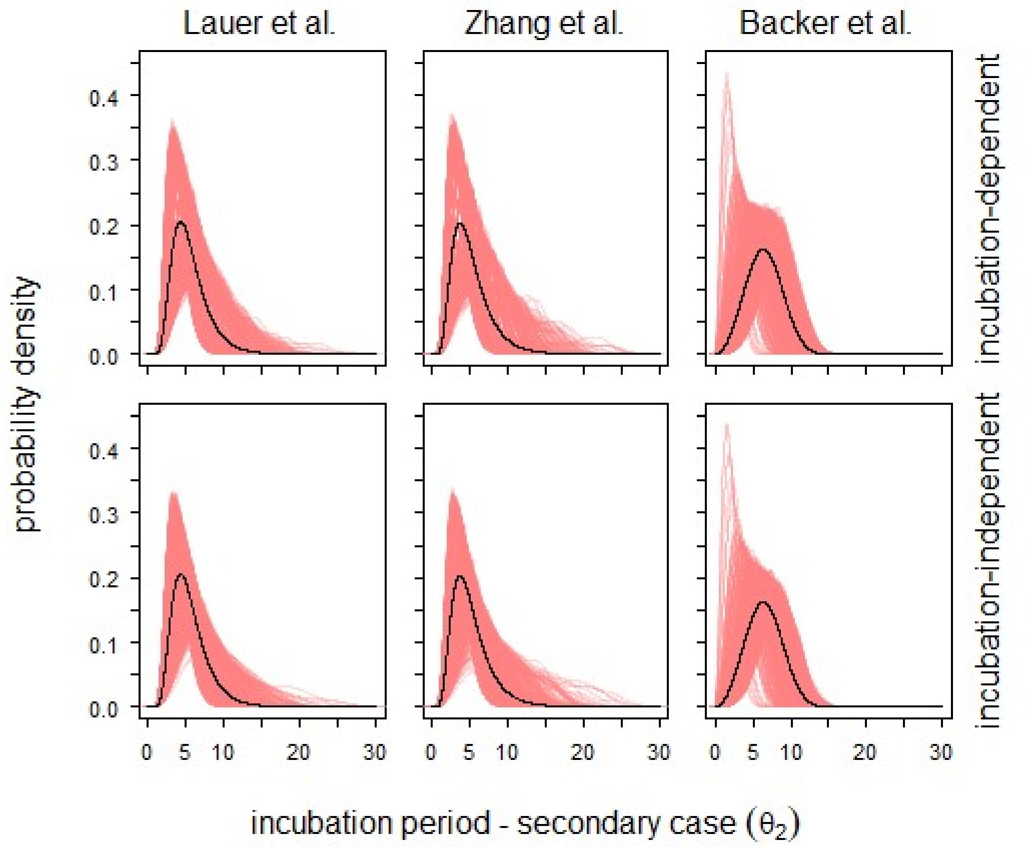


**Fig. S4** Overlaid posterior distributions for the incubation period of the secondary case ($\theta_2$) of each case pair in the post-lockdown period. Black lines show the incubation period prior for each analysis.
