## Supplementary figures and images for "Transmission of SARS-CoV-2 before and after symptom onset: impact of nonpharmaceutical interventions in China"

### Fig S5

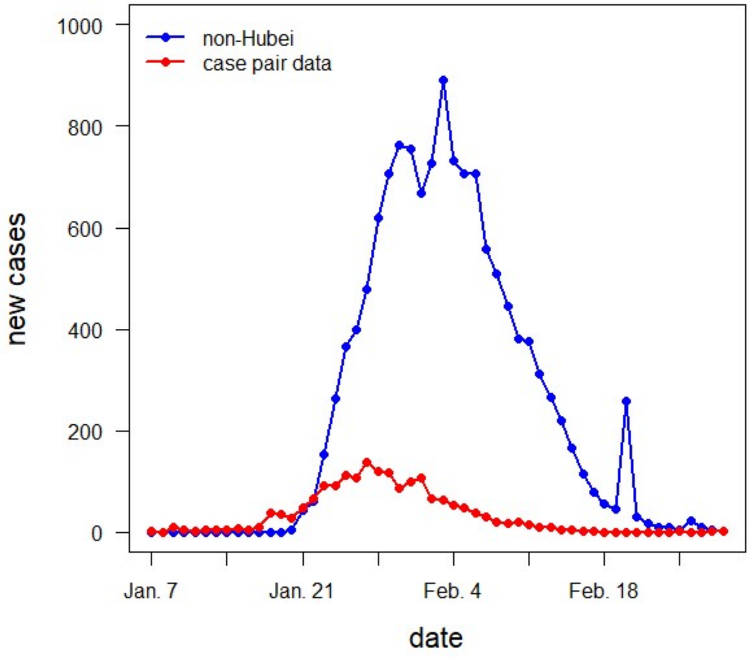


**Fig. S5** Case incidence based on case pairs (red) and non-Hubei cases (blue).
