## Supplementary material for "Transmission of SARS-CoV-2 before and after symptom onset: impact of nonpharmaceutical interventions in China": Fig S6

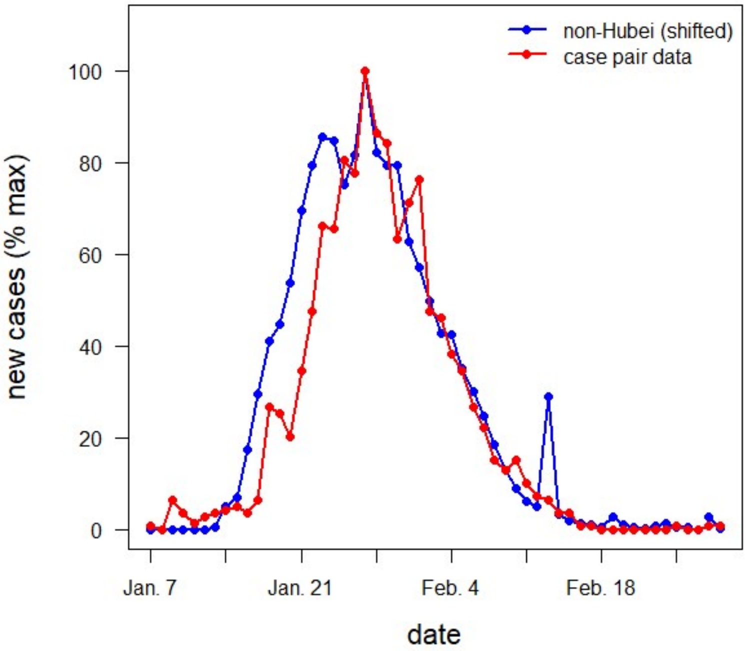


**Fig. S6** Case incidence based on case pairs (red) and non-Hubei cases (blue), with both curves scaled to their respective maximums and the non-Hubei incidence curve shifted 7 days to the left.
