## Supplementary material for "Transmission of SARS-CoV-2 before and after symptom onset: impact of nonpharmaceutical interventions in China": Fig S7

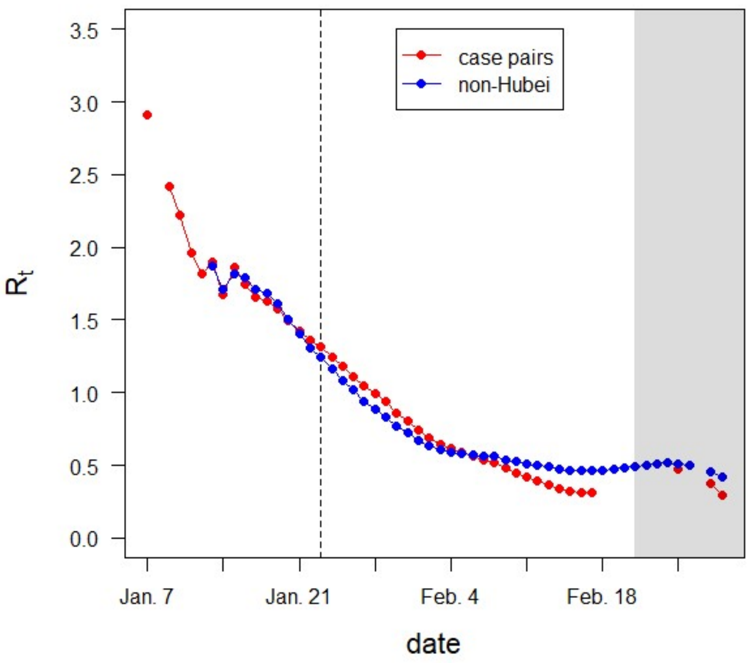


**Fig. S7** Daily estimates of $R_t$ based on case pair incidence data (red) and non-Hubei incidence data (blue). Dashed vertical line, start of initial lockdown in Wuhan on Jan. 23; gray shading, region in which $R_t$ is likely to be underestimated due to right-truncation.
