## Supplementary material for "Transmission of SARS-CoV-2 before and after symptom onset: impact of nonpharmaceutical interventions in China": Fig S8

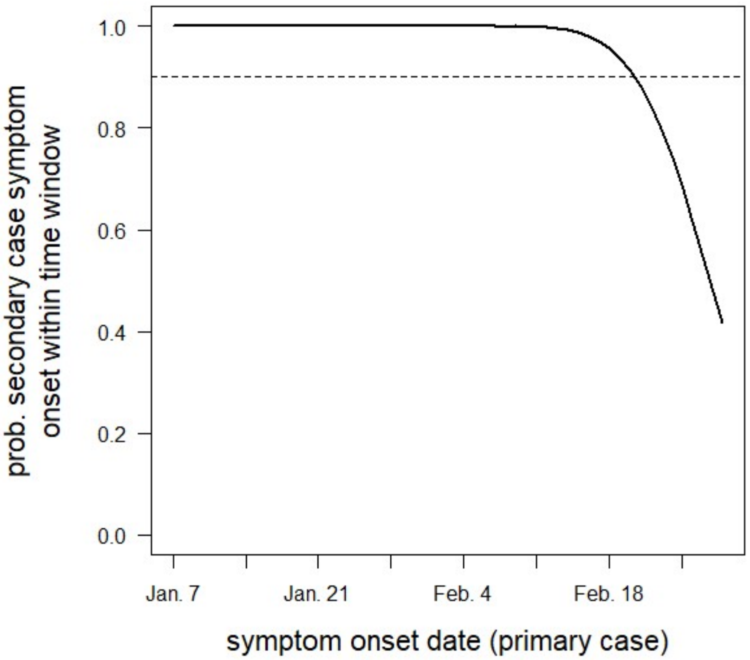


**Fig. S8** Probability of a secondary case developing symptoms by $t_{\max}$ (Feb. 29) vs. time of primary case symptom onset. Dashed line shows probability cutoff (0.9).
