## Supplementary material for "Transmission of SARS-CoV-2 before and after symptom onset: impact of nonpharmaceutical interventions in China": S1 Text

**Applying deviance information criteria (DIC) to models of pre-symptomatic transmission of SARS-CoV-2**

A comprehensive guide to deviance information criteria (DIC) for missing data models exists in the paper by Celeux et al; here, we introduce only those definitions and concepts applicable to our models. The first part is devoted to DIC in general, while the second part shows how DIC is calculated for our models in particular.

<u>*Part I: Theory*</u>

*Introduction*

Let $f(\mathbf{y}|\Theta)$ be the likelihood of observing some data $\mathbf{y}$, given a model $f$ with parameter(s) $\Theta$.

The *deviance* of model $f$ with parameters $\Theta$ is defined as

$$D(\Theta) = -2\log f(\mathbf{y}|\Theta) + 2\log h(\mathbf{y})$$

where $h(\mathbf{y})$ is a function of the data alone, i.e. it does not depend on $\Theta$.

The deviance information criterion, or DIC, is defined by the following expression:

$$\text{DIC} = \overline{D(\Theta)} + p_D$$

$\overline{D(\Theta)}$ is the posterior mean deviance, a Bayesian measure of model fit, which is found by taking the expectation of $D(\Theta)$ over the posterior distribution of $\Theta$:

$$\overline{D(\Theta)} = E_\Theta[-2\log f(\mathbf{y}|\Theta)|\mathbf{y}] + 2\log h(\mathbf{y})$$

$p_D$ is the effective number of parameters, a measure of model complexity, and is given by

$$p_D = \overline{D(\Theta)} - D(\tilde{\Theta})$$

where $\tilde{\Theta}$ is an estimate for $\Theta$ given $\mathbf{y}$, such as the posterior mean $E_\Theta[\Theta|\mathbf{y}]$.

We can therefore rewrite DIC as follows:

$$\text{DIC} = 2\overline{D(\Theta)} - D(\tilde{\Theta})$$

$$= -4E_\Theta[\log f(\mathbf{y}|\Theta)|\mathbf{y}] + 2\log f(\mathbf{y}|\tilde{\Theta}) + 2\log h(\mathbf{y})$$

Since $h(\mathbf{y})$ does not depend on the model or parameters, it is irrelevant for model comparison; we can therefore set $h(\mathbf{y}) = 1$ which makes $2\log h(\mathbf{y}) = 0$, simplifying the expression for DIC to

$$\text{DIC} = -4E_\Theta[\log f(\mathbf{y}|\Theta)|\mathbf{y}] + 2\log f(\mathbf{y}|\tilde{\Theta})$$

*Estimating DIC from Markov chain Monte Carlo (MCMC)*

DIC can be readily applied to model fitting by MCMC because the posterior distribution of $\Theta$ is approximated by the post-convergence Markov chain. As long as the likelihood function $f(\mathbf{y}|\Theta)$ is available in closed form, the posterior mean deviance, $\overline{D(\Theta)}$, can be estimated by averaging $D(\Theta)$ over all of the steps in the chain, and $D(\tilde{\Theta})$ simply requires an estimator of $\Theta$, such as the posterior mean.

*Extension of DIC to missing data models*

With data-augmented models, the likelihood function $f(\mathbf{y}|\Theta)$ is often not available in closed form, because it depends on missing data $\mathbf{z}$ as well as the observed data $\mathbf{y}$. In this case, $f(\mathbf{y}|\Theta)$ is called as the *observed* likelihood, while $f(\mathbf{y},\mathbf{z}|\Theta)$ is termed the *complete* likelihood.

The DIC for a missing data model can therefore be re-written in terms of the complete likelihood, as follows:

$$\text{DIC} = -4E_{\Theta}[\log f(\mathbf{y},\mathbf{z}|\Theta)|\mathbf{y},\mathbf{z}] + 2\log f(\mathbf{y},\mathbf{z}|E_{\Theta}[\Theta|\mathbf{y},\mathbf{z}])$$

Since the data $\mathbf{z}$ are, by definition, missing, this quantity can not be computed directly; however, if the distribution of $\mathbf{Z}$ is known or can be approximated (e.g. using a data augmentation MCMC algorithm), it is sufficient to take the expectation of DIC with respect to $\mathbf{Z}$:

$$\text{DIC} = E_{\mathbf{Z}}[\text{DIC}(\mathbf{y},\mathbf{Z})|\mathbf{y}]$$

$$= -4E_{\Theta,\mathbf{Z}}[\log f(\mathbf{y},\mathbf{Z}|\Theta)|\mathbf{y}] + 2E_{\mathbf{Z}}[\log f(\mathbf{y},\mathbf{Z}|E_{\Theta}[\Theta|\mathbf{y},\mathbf{Z}])|\mathbf{y}]$$

The first term in this expression can be estimated using the posterior distributions of $\Theta$ and $\mathbf{Z}$ from a data augmentation MCMC algorithm, but the second term requires calculation of the posterior mean $E_{\Theta}[\Theta|\mathbf{y},\mathbf{Z}]$ for each value of $\mathbf{Z}$, which is inconvenient. However, this term can be reformulated to make estimation more straightforward.
Recall that the formula for DIC is as follows:

$$\text{DIC} = 2\overline{D(\Theta)} - D(\tilde{\Theta})$$

And recall that, if we set $h(\mathbf{y}) = 1$, the formula for deviance simplifies to

$$D(\Theta) = -2\log f(\mathbf{y}|\Theta)$$

and therefore the second term in the DIC expression can be rewritten as

$$D(\tilde{\Theta}) = -2\log f(\mathbf{y}|\tilde{\Theta})$$

Expectation-maximization (EM) algorithms suggest a way to approximate the log-likelihood in the context of missing data:

$$\log f(\mathbf{y}|\Theta) = E_{\mathbf{Z}}[\log f(\mathbf{y},\mathbf{Z}|\hat{\Theta}(\mathbf{y}))|\mathbf{y},\hat{\Theta}(\mathbf{y})]$$

where $\hat{\Theta}(\mathbf{y})$ is an estimator of $\Theta$ based on the observed data $\mathbf{y}$.

Thus, an alternative way of writing DIC is as follows:

$$\text{DIC} = -4E_{\Theta,\mathbf{Z}}[\log f(\mathbf{y},\mathbf{Z}|\Theta)|\mathbf{y}] + 2E_{\mathbf{Z}}[\log f(\mathbf{y},\mathbf{Z}|\hat{\Theta}(\mathbf{y}))|\mathbf{y},\hat{\Theta}(\mathbf{y})]$$

The second term in this expression can be estimated by running a second MCMC with the parameters $\Theta$ fixed at $\hat{\Theta}(\mathbf{y})$ and taking the expectation of the log-likelihood with respect to the posterior distribution of the missing data $\mathbf{Z}$.

*Part II: Application*

We now show how DIC is calculated for the models of pre-symptomatic transmission described in the Materials & Methods. Recall that the generation interval $\tau$ is assumed to follow a gamma distribution with shape parameter $\alpha$ and rate parameter $\beta$ (incubation-independent model) or $\beta/\theta_1$ (incubation-dependent model). The incubation periods of the infector and infectee are denoted $\theta_1$ and $\theta_2$, respectively, while $\delta$ refers to the serial interval.

We used an MCMC algorithm with data augmentation to estimate the parameters $\alpha$ and $\beta$, as well as the missing data $\boldsymbol{\theta_1}$ and $\boldsymbol{\theta_2}$, by fitting to the serial interval data $\boldsymbol{\delta}$. Thus, in the notation of Part 1, the serial interval data $\boldsymbol{\delta}$ are the observed data $\mathbf{y}$, the incubation periods $\boldsymbol{\theta_1}$ and $\boldsymbol{\theta_2}$ comprise the missing data $\mathbf{z}$, and the parameters $\alpha$ and $\beta$ are the model parameters $\Theta$.

At the end of Part 1, we arrived at the following expression for DIC for a missing data model:

$$\text{DIC} = -4E_{\Theta,\mathbf{Z}}[\log f(\mathbf{y},\mathbf{Z}|\Theta)|\mathbf{y}] + 2E_{\mathbf{Z}}[\log f(\mathbf{y},\mathbf{Z}|\hat{\Theta}(\mathbf{y}))|\mathbf{y},\hat{\Theta}(\mathbf{y})]$$

Rewriting this for our models, we get:

$$\text{DIC} = -4E_{\alpha,\beta,\boldsymbol{\theta_1},\boldsymbol{\theta_2}}[\log f(\boldsymbol{\delta},\boldsymbol{\theta_1},\boldsymbol{\theta_2}|\alpha,\beta)|\boldsymbol{\delta}] + 2E_{\boldsymbol{\theta_1},\boldsymbol{\theta_2}}[\log f(\boldsymbol{\delta},\boldsymbol{\theta_1},\boldsymbol{\theta_2}|\hat{\alpha},\hat{\beta})|\boldsymbol{\delta},\hat{\alpha},\hat{\beta}]$$

From this point on, since our focus is application rather than theory, we drop several redundant conditionals from the equations to improve readability. This simplifies this above to

$$\text{DIC} = -4E_{\alpha,\beta,\boldsymbol{\theta_1},\boldsymbol{\theta_2}}[\log f(\boldsymbol{\delta},\boldsymbol{\theta_1},\boldsymbol{\theta_2}|\alpha,\beta)] + 2E_{\boldsymbol{\theta_1},\boldsymbol{\theta_2}}[\log f(\boldsymbol{\delta},\boldsymbol{\theta_1},\boldsymbol{\theta_2}|\hat{\alpha},\hat{\beta})]$$

The first component of this expression can be rewritten as follows:

$$-4E_{\alpha,\beta,\boldsymbol{\theta_1},\boldsymbol{\theta_2}}[\log f(\boldsymbol{\delta},\boldsymbol{\theta_1},\boldsymbol{\theta_2}|\alpha,\beta)]$$

$$= -\frac{4}{M}\sum_{i=1}^{M}\log f(\boldsymbol{\delta},\boldsymbol{\theta_1}^{(i)},\boldsymbol{\theta_2}^{(i)}|\alpha^{(i)},\beta^{(i)})$$

$$= -\frac{4}{M}\sum_{i=1}^{M}\sum_{k=1}^{N}\log f(\delta_k,\theta_{1(k)}^{(i)},\theta_{2(k)}^{(i)}|\alpha^{(i)},\beta^{(i)})$$

$$= -\frac{4}{M}\sum_{i=1}^{M}\sum_{k=1}^{N}\log f(\delta_k|\theta_{1(k)}^{(i)},\theta_{2(k)}^{(i)},\alpha^{(i)},\beta^{(i)}) + \log f_\theta(\theta_{1(k)}^{(i)}) + \log f_\theta(\theta_{2(k)}^{(i)})$$

$$= -\frac{4}{M}\sum_{i=1}^{M}\sum_{k=1}^{N} \log f_\tau(\delta_k + \theta_{1(k)}^{(i)} - \theta_{2(k)}^{(i)}|\alpha^{(i)}, \beta^{(i)}) + \log f_\theta(\theta_{1(k)}^{(i)}) + \log f_\theta(\theta_{2(k)}^{(i)})$$

where $M$ is the number of iterations in the thinned converged Markov chain, with a superscript $^{(i)}$ denoting the $i$th iteration; $N$ is the number of infector-infectee pairs represented in the serial interval data set, with a subscript $_{(k)}$ denoting the $k$th pair; $f_\tau$ is the generation interval distribution (with parameters $\alpha$ and $\beta$); and $f_\theta$ is the prior for the incubation period.

The second component of the expression can similarly be rewritten:

$$2E_{\boldsymbol{\theta}_1,\boldsymbol{\theta}_2}[\log f(\boldsymbol{\delta}, \boldsymbol{\theta}_1, \boldsymbol{\theta}_2|\hat{\alpha}, \hat{\beta})]$$

$$= \frac{2}{M'}\sum_{i=1}^{M'} \log f(\boldsymbol{\delta}, {\boldsymbol{\theta}_1}'^{(i)}, {\boldsymbol{\theta}_2}'^{(i)}|\hat{\alpha}, \hat{\beta})$$

$$= \frac{2}{M'}\sum_{i=1}^{M'}\sum_{k=1}^{N} \log f(\delta_k, {\theta_{1(k)}}'^{(i)}, {\theta_{2(k)}}'^{(i)}|\hat{\alpha}, \hat{\beta})$$

$$= \frac{2}{M'}\sum_{i=1}^{M'}\sum_{k=1}^{N} \log f(\delta_k|{\theta_{1(k)}}'^{(i)}, {\theta_{2(k)}}'^{(i)}, \hat{\alpha}, \hat{\beta}) + \log f_\theta({\theta_{1(k)}}'^{(i)}) + \log f_\theta({\theta_{2(k)}}'^{(i)})$$

$$= \frac{2}{M'}\sum_{i=1}^{M'}\sum_{k=1}^{N} \log f_\tau(\delta_k + {\theta_{1(k)}}'^{(i)} - {\theta_{2(k)}}'^{(i)}|\hat{\alpha}, \hat{\beta}) + \log f_\theta({\theta_{1(k)}}'^{(i)}) + \log f_\theta({\theta_{2(k)}}'^{(i)})$$

where $\hat{\alpha}$ and $\hat{\beta}$ are the parameter estimates (in this case, posterior means) from the original MCMC, and apostrophes indicate quantities from a secondary MCMC in which $\alpha$ and $\beta$ are fixed at $\hat{\alpha}$ and $\hat{\beta}$, respectively.
