## Supplementary material for "Transmission of SARS-CoV-2 before and after symptom onset: impact of nonpharmaceutical interventions in China": Table S1

| | Primary infection location | | | | |
|---|---|---|---|---|---|
| **Secondary infection location** | | **Hubei** | **Non-Hubei** | **Unknown** | **Marginal** |
| | **Hubei** | 62 (7.1%) | 3 (0.3%) | 1 (0.1%) | 66 (7.6%) |
| | **Non-Hubei** | 324 (37.1%) | 356 (40.8%) | 55 (6.3%) | 735 (84.2%) |
| | **Unknown** | 11 (1.3%) | 8 (0.9%) | 53 (6.1%) | 72 (8.2%) |
| | **Marginal** | 397 (45.5%) | 367 (42.0%) | 109 (12.5%) | 873 (100%) |

**Table S1** Chinese provinces in which primary and secondary cases were presumed to be infected.
