## Supplementary material for "Transmission of SARS-CoV-2 before and after symptom onset: impact of nonpharmaceutical interventions in China": Table S2

| Parameter | Prior | Source |
|---|---|---|
| $\alpha$ | Uniform on [0,100] | n/a |
| $\beta$ | Uniform on [0,100] | n/a |
| $\theta$ | Lognormal | Lauer et al. (*1*) |
| $\theta$ | Lognormal | Zhang et al. (*2*) |
| $\theta$ | Weibull | Backer et al. (*3*) |

**Table S2** Prior distributions for MCMC parameter estimation.

1. S. A. Lauer, K. H. Grantz, Q. Bi, F. K. Jones, Q. Zheng, H. R. Meredith, A. S. Azman, N. G. Reich, J. Lessler, The Incubation Period of Coronavirus Disease 2019 (COVID-19) From Publicly Reported Confirmed Cases: Estimation and Application. *Ann Intern Med* **172**, 577-582 (2020).
2. J. Zhang, M. Litvinova, W. Wang, Y. Wang, X. Deng, X. Chen, M. Li, W. Zheng, L. Yi, X. Chen, Q. Wu, Y. Liang, X. Wang, J. Yang, K. Sun, I. M. Longini, Jr., M. E. Halloran, P. Wu, B. J. Cowling, S. Merler, C. Viboud, A. Vespignani, M. Ajelli, H. Yu, Evolving epidemiology and transmission dynamics of coronavirus disease 2019 outside Hubei province, China: a descriptive and modelling study. *Lancet Infect Dis* 10.1016/S1473-3099(20)30230-9 (2020).
3. J. A. Backer, D. Klinkenberg, J. Wallinga, Incubation period of 2019 novel coronavirus (2019-nCoV) infections among travellers from Wuhan, China, 20-28 January 2020. *Euro Surveill* **25**(2020).
