## Supplementary material for "Transmission of SARS-CoV-2 before and after symptom onset: impact of nonpharmaceutical interventions in China": Table S3

| Time period | Generation interval model | Incubation period prior | | | | | DIC |
|---|---|---|---|---|---|---|---|
| | | | mean | 95% CI | mean | 95% CI | |
| Pre-lockdown | incubation-independent | Lauer et al. | 3.71 | (2.57, 4.99) | 0.495 | (0.344, 0.666) | 2899 |
| | | Zhang et al. | 4.05 | (2.66, 6.68) | 0.541 | (0.355, 0.886) | 2916 |
| | | Backer et al. | 3.99 | (2.72, 5.71) | 0.527 | (0.362, 0.745) | 2983 |
| | incubation-dependent | Lauer et al. | 2.87 | (2.12, 3.83) | 2.09 | (1.49, 2.86) | 2932 |
| | | Zhang et al. | 2.98 | (2.23, 3.87) | 2.04 | (1.48, 2.73) | 2955 |
| | | Backer et al. | 3.03 | (2.22, 4.00) | 2.58 | (1.85, 3.41) | 3005 |
| Post-lockdown | incubation-independent | Lauer et al. | 1.55 | (1.27, 1.91) | 0.398 | (0.325, 0.490) | 8896 |
| | | Zhang et al. | 1.65 | (1.31, 2.06) | 0.430 | (0.338, 0.543) | 8923 |
| | | Backer et al. | 1.51 | (1.23, 1.82) | 0.382 | (0.313, 0.459) | 9190 |
| | incubation-dependent | Lauer et al. | 1.25 | (1.05, 1.49) | 1.73 | (1.43, 2.09) | 8971 |
| | | Zhang et al. | 1.34 | (1.12, 1.60) | 1.77 | (1.43, 2.15) | 9014 |
| | | Backer et al. | 1.24 | (1.05, 1.45) | 1.99 | (1.66, 2.37) | 9222 |

**Table S3** Posterior means and 95% credible intervals for the parameters of the generation interval distribution, plus deviance information criterion (DIC) estimates for each model and each time period.
