## Supplementary material for "Transmission of SARS-CoV-2 before and after symptom onset: impact of nonpharmaceutical interventions in China": Table S4

| Time period | Generation interval model | Incubation period prior | mean | | sd | |
|---|---|---|---|---|---|---|
| | | | *mean* | *95% CI* | *mean* | *95% CI* |
| Pre-lockdown | incubation-independent | Lauer et al. | 7.50 | (6.81, 8.20) | 3.95 | (3.32, 4.74) |
| | | Zhang et al. | 7.49 | (6.80, 8.22) | 3.83 | (2.90, 4.65) |
| | | Backer et al. | 7.58 | (6.90, 8.28) | 3.86 | (3.18, 4.61) |
| | incubation-dependent | Lauer et al. | 7.63 | (6.89, 8.42) | 5.99 | (5.14, 6.97) |
| | | Zhang et al. | 7.62 | (6.87, 8.45) | 6.28 | (5.41, 7.24) |
| | | Backer et al. | 7.63 | (6.94, 8.38) | 5.48 | (4.76, 6.34) |
| Post-lockdown | incubation-independent | Lauer et al. | 3.90 | (3.59, 4.24) | 3.15 | (2.78, 3.53) |
| | | Zhang et al. | 3.84 | (3.49, 4.18) | 3.01 | (2.60, 3.43) |
| | | Backer et al. | 3.95 | (3.63, 4.30) | 3.23 | (2.89, 3.61) |
| | incubation-dependent | Lauer et al. | 3.99 | (3.66, 4.34) | 4.28 | (3.83, 4.77) |
| | | Zhang et al. | 3.96 | (3.62, 4.32) | 4.31 | (3.84, 4.85) |
| | | Backer et al. | 4.04 | (3.71, 4.38) | 4.14 | (3.73, 4.59) |

**Table S4** Posterior means and 95% credible intervals for the mean and standard deviation of the generation interval distribution.
