## Supplementary material for "Transmission of SARS-CoV-2 before and after symptom onset: impact of nonpharmaceutical interventions in China": Table S5

| Generation interval model | Incubation period prior | Pre-lockdown | | Post-lockdown | |
|---|---|---|---|---|---|
| | | *mean* | *95% CI* | *mean* | *95% CI* |
| incubation-independent | Lauer et al. | 34.4% | (28.3%, 41.3%) | 71.0% | (67.6%, 74.2%) |
| | Zhang et al. | 30.7% | (24.0%, 37.7%) | 68.1% | (64.5%, 71.7%) |
| | Backer et al. | 43.7% | (37.1%, 50.4%) | 77.5% | (74.5%, 80.3%) |
| incubation-dependent | Lauer et al. | 37.8% | (31.4%, 44.5%) | 75.1% | (71.8%, 78.3%) |
| | Zhang et al. | 34.1% | (27.7%, 40.8%) | 73.2% | (69.5%, 76.8%) |
| | Backer et al. | 47.0% | (40.2%, 53.7%) | 80.6% | (77.6%, 83.5%) |

**Table S5** Posterior means and 95% credible interavls for the relative frequency of presymptomatic transmission during the pre-lockdown and post-lockdown periods.
