## Supplementary material for "Transmission of SARS-CoV-2 before and after symptom onset: impact of nonpharmaceutical interventions in China": Table S6

| Generation interval model | Incubation period prior | presymptomatic transmission | | transmission post-symptom onset | |
|---|---|---|---|---|---|
| | | *mean* | *95% CI* | *mean* | *95% CI* |
| incubation-independent | Lauer et al. | -15.5% | (-30.6%, +2.56%) | -82.0% | (-84.5%, -79.0%) |
| | Zhang et al. | -8.62% | (-27.0%, +16.3%) | -81.3% | (-84.0%, -78.3%) |
| | Backer et al. | -27.5% | (-37.9%, -15.1%) | -83.7% | (-86.3, -80.7%) |
| incubation-dependent | Lauer et al. | -18.7% | (-31.8%, -2.11%) | -83.7% | (-86.3%, -80.7%) |
| | Zhang et al. | -12.0% | (-27.6%, +7.51%) | -83.5% | (-86.1%, -80.5%) |
| | Backer et al. | -29.9% | (-39.4%, -18.2%) | -85.1% | (-87.9%, -81.9%) |

**Table S6** Posterior means and 95% credible intervals for the % change in the absolute frequency of presymptomatic transmission and transmission post-symptom onset after the introduction of NPIs.
