## Supplementary material for "Transmission of SARS-CoV-2 before and after symptom onset: impact of nonpharmaceutical interventions in China": Table S7

| Transmission type | Pre-lockdown | Post-lockdown |
|---|---|---|
| Household | 7 | 35 |
| Non-household | 26 | 53 |
| Family | 81 | 365 |
| Non-family | 71 | 143 |
| Imported | 126 | 303 |
| Non-imported | 61 | 292 |

**Table S7** Numbers of six types of transmission events: attributable to household contact; attributable to non-household contact; between family members; between non-family members; imported (primary and secondary cases infected in different cities); non-imported (primary and secondary cases infected in the same city). Contact type, relationship, and infection location data were not available for all case pairs, so the totals for household/non-household (121), family/non-family (660), and imported/non-imported (782) are different from one another and less than the total number of case pairs (873).
